# Treating Vision, Not Signs: A Post-hoc Analysis Evaluating BCVA as an Early Indicator in Treat-and-Extend Management of DME

**DOI:** 10.1101/2025.07.13.25331469

**Authors:** Aly Hamza Khowaja, Kholood Janjua, Haider Ali, Rubbia Afridi, Zoha Zahid Fazal, Mohammad Abdul Saqhlain Shaik, Mohamed Ibrahim Ahmed, Muhammad Sohail Halim, Theodore Leng, Carolyn K. Pan, Quan Dong Nguyen, Yasir Jamal Sepah

**Affiliations:** Medical College, The Aga Khan University Hospital, Karachi, Sindh, Pakistan; Ophthalmology, Stanford University Medicine, Palo Alto, California, United States; Ocular Imaging Research and Reading Center, Sunnyvale, California, United States

**Keywords:** ocular biomarkers, precision medicine, OCT, Central Retinal Thickness, anti-VEGF

## Abstract

**Purpose:** To determine whether significant changes in best-corrected visual acuity (BCVA) precede or coincide with increases in central retinal thickness (CRT) in diabetic macular edema (DME) during a treat-and-extend (T&E) regimen following initial edema resolution.

**Methods:** This post-hoc analysis included 60 eyes (60 participants) from the READ-3 clinical trial that achieved CRT <250 µm and were followed until edema recurrence. Following a six-month ranibizumab loading phase, patients were monitored through 24 months with as-needed retreatment. Significant changes were defined as ≥4 Early Treatment Diabetic Retinopathy Study (ETDRS) letters and ≥30 µm on time-domain optical coherence tomography (TD-OCT). The temporal relationship between functional (BCVA) and anatomical (CRT) changes was analyzed.

**Results:** Median time to edema resolution was 10 months (IQR: 6-16) and to recurrence was 3 months (IQR: 2-3). 52 eyes (86.7%) had functional worsening and 43 (71.7%) had anatomical worsening. In 39 eyes exhibiting both types of deterioration, changes were concurrent in 24 (61.5%). Vision loss preceded anatomical recurrence (BCVA-led) in 23.1% of eyes, with a lead time of 1-4 months. Conversely, anatomical thickening preceded vision loss (OCT-led) in 15.4% of eyes, by a maximum of 2 months.

**Conclusions:** BCVA fluctuations frequently mirror CRT changes and can precede structural relapse, suggesting that BCVA is a sensitive indicator of DME activity. In resource-limited settings, BCVA may allow for earlier detection of recurrence than OCT alone.

**Translational Relevance:** Functional vision loss can precede structural edema recurrence, supporting the potential for home-based BCVA monitoring as a validated bridge for timely clinical intervention in DME.

## Introduction

Diabetic macular edema (DME) is a leading cause of preventable vision loss among working-age individuals with diabetic retinopathy (DR)^1^. The advent of intravitreal anti-vascular endothelial growth factor (anti-VEGF) therapy has revolutionized DME management, yielding significant visual gains in clinical trials. Historically, regulatory approval and trial endpoints have centered on functional improvements, specifically best-corrected visual acuity (BCVA) gains^2,3^. However, in routine practice ophthalmologists often rely on optical coherence tomography (OCT) findings, such as changes in central retinal thickness (CRT) or central subfield thickness, to guide re-treatment. This shift toward anatomical endpoints reflects the assumption that retinal thickening correlates with, and even precedes, vision loss^4^.

In reality, the structure-function relationship in DME is complex and variable. Not all patients experience parallel improvements in BCVA and CRT. Real-world data suggest that fewer than 25% of eyes achieve both a dry macula and BCVA 20/40 after nine anti-VEGF injections^5^. Such misalignment means some patients have persistent vision impairment despite resolved edema, while others maintain good vision despite residual edema. If physicians treat based solely on OCT abnormalities, patients might receive injections even when functional vision is stable. This discordance can lead to overly aggressive treatment schedules and increases the burden of frequent clinic visits and injections on patients and healthcare systems. Conversely, monitoring primarily by OCT may delay detection of functional decline in cases where BCVA worsens before obvious OCT changes. In such scenarios, insisting on costly, monthly OCT monitoring for every patient may become impractical.

While the technological precision of OCT is undeniable, the current over-reliance on anatomical endpoints risks distancing clinicians from patients’ primary concern: vision. By recentering management around visual acuity, we align treatment triggers with the patient’s lived experience rather than treating isolated tomographic signs. There is thus a pressing need for practical monitoring solutions that can detect disease activity as a timely and patient-centric alternative without over-reliance on clinic-based OCT. We hypothesize that clinically meaningful changes in BCVA (≥4 ETDRS letters) might serve as an early indicator of DME activity, occurring concurrently with or even ahead of CRT changes, including in eyes that have been successfully treated to resolution of edema. To explore this hypothesis, we performed a post-hoc analysis of the READ-3 (Ranibizumab for Edema of the mAcula in Diabetes, Protocol 3) trial^6^ – a prospective, multicenter DME study with high-frequency OCT and BCVA follow-up. By analyzing the lag between functional and anatomical changes during a treat-and-extend protocol, we tested whether clinically meaningful BCVA changes (≥4 ETDRS letters) precede, coincide with, or follow OCT-detected CRT increases after initial macular drying, which has implications for monitoring strategies and resource allocation. We also considered the implications for a home-based BCVA monitoring system that could complement or substitute for OCT in routine DME management.

## Methods

### Study Design and Population

This is a post-hoc analysis of READ-3, a 24-month randomized clinical trial conducted at 13 U.S. sites. The trial enrolled adults with DME meeting typical inclusion criteria, including baseline CST ≥250 µm on time-domain OCT and BCVA 20/40 to 20/320 (ETDRS letters). Key exclusion criteria were recent macular laser or intraocular steroid, or anti-VEGF treatment within 2–3 months. Each patient contributed one study eye; if both were eligible, the eye with greater CRT was selected. All participants provided informed consent, and the study adhered to the Declaration of Helsinki and was registered on clinicaltrials.gov (NCT01077401)^7^.

### Treatment Protocol

Enrolled eyes were randomized 1:1 to monthly intravitreal ranibizumab 0.5 mg or 2.0 mg for six consecutive months (baseline through Month 5). The month 6 visit, which was also the primary endpoint of the trial occurred 4 weeks after the sixth injection.^6^ At and beyond Month 6, treat-and-extend management was implemented: patients were evaluated monthly and received additional ranibizumab injections only if OCT showed recurrent macular edema (central subfield thickness ≥250 µm on time-domain OCT (TD-OCT), or any macular fluid detected on time-domain or spectral-domain OCT). If no edema was present, the injection could be deferred, and the patient examined again the following month. (Note: This regimen is effectively a pro re nata (PRN, as-needed) approach with monthly monitoring; for simplicity we refer to it as “treat-and-extend,” although no fixed extension interval was predetermined beyond monthly evaluations.) All study visits included BCVA measurement (following ETDRS protocol^8^ by certified examiners), OCT imaging, and ophthalmic examinations.

### Study Cohort and Inclusion Criteria

We identified eyes that achieved complete resolution of edema at any visit after the mandatory monthly loading dose visits, defined as CRT <250 µm with no intraretinal or subretinal fluid visible on OCT. For this analysis, the first occurrence of an “edema-free” retina, as well as documented evidence of deferred injection, was considered the start-point, or the eligibility visit, for each eye. Included eyes were followed from this visit to the recurrence of edema and retreatment. Eyes that (1) did not achieve edema resolution; (2) did not have edema recurrence following treatment; (3) were treated pro-actively before evidence of recurrence, were all excluded from analysis.

### Outcome Measures

For the present analysis, we tracked two main outcomes: (1) Lag between functional and anatomical changes, specifically, whether BCVA changes occurred before, at the same time, or after corresponding CRT changes; and (2) Injection-free interval: the duration each eye remained in the “treat-and-extend” phase of deferred re-injection after eligibility, which reflects how long edema stayed in remission.

### Definitions

We defined the eligibility visit as the first visit for each patient that saw the concurrence of CRT <250 µm with no intraretinal or subretinal fluid visible on TD-OCT and deferral of injection. By protocol definition, this could only occur after completion of the initial loading phase of mandatory injections. A clinically significant BCVA change was defined as a gain or loss of ≥4 letters from the previous visit (approximately equivalent to ≥0.08 logMAR change), based on test-retest variability of ETDRS charts and prior definitions of meaningful change.^9^ A significant CRT change was defined as an increase or decrease of >30 µm on TD-OCT, consistent with thresholds used in DME studies for meaningful edema fluctuation. A BCVA–CRT or ‘B-C’ lag was said to occur if a significant change in BCVA was not observed in the *same* visit as a significant change in CRT for that eye. In other words, if one parameter changed at visit *n* and the other changed at a later visit *n+x* (or had changed earlier at *n–x*), we considered a lag of *x* months to be present. By this definition, we classify as BCVA-leading cases those where BCVA change came first, CRT-leading cases those where CRT changed first, and concurrent cases those where both changed in the same month (lag = 0). Eyes that only met the criteria for one parameter (missing either significant BCVA or CRT worsening during follow-up, but not both) were tracked as well.

### Data Analysis

We summarized baseline patient characteristics and the distribution of lag categories using descriptive statistics. For each eye, the timing (months) of the first occurrence of significant functional change (≥4-letter drop in BCVA) and significant anatomical change (≥30 µm rise in CRT) during the eligible study interval was recorded. Eyes were then categorized by the temporal sequence of these events: BCVA change preceding CRT change, simultaneous change, CRT change preceding BCVA change, or no significant event. All statistical tests were 2-tailed with a significance level of α=0.05. Analyses were performed using Stata 15.1 (StataCorp, College Station, TX).

## Results

### Cohort Identification

Of the 155 eyes in the READ-3 trial, 81 eyes reached at least one edema-free event. 21 of these eyes had re-injection before any change in BCVA or any change on OCT could be observed. The 60 eyes with changes in at least one parameter were hence identified. A further subset of 39 eyes with changes on both, OCT and BCVA, was also identified. The process is depicted as **Figure 1**.

**Figure 1.**
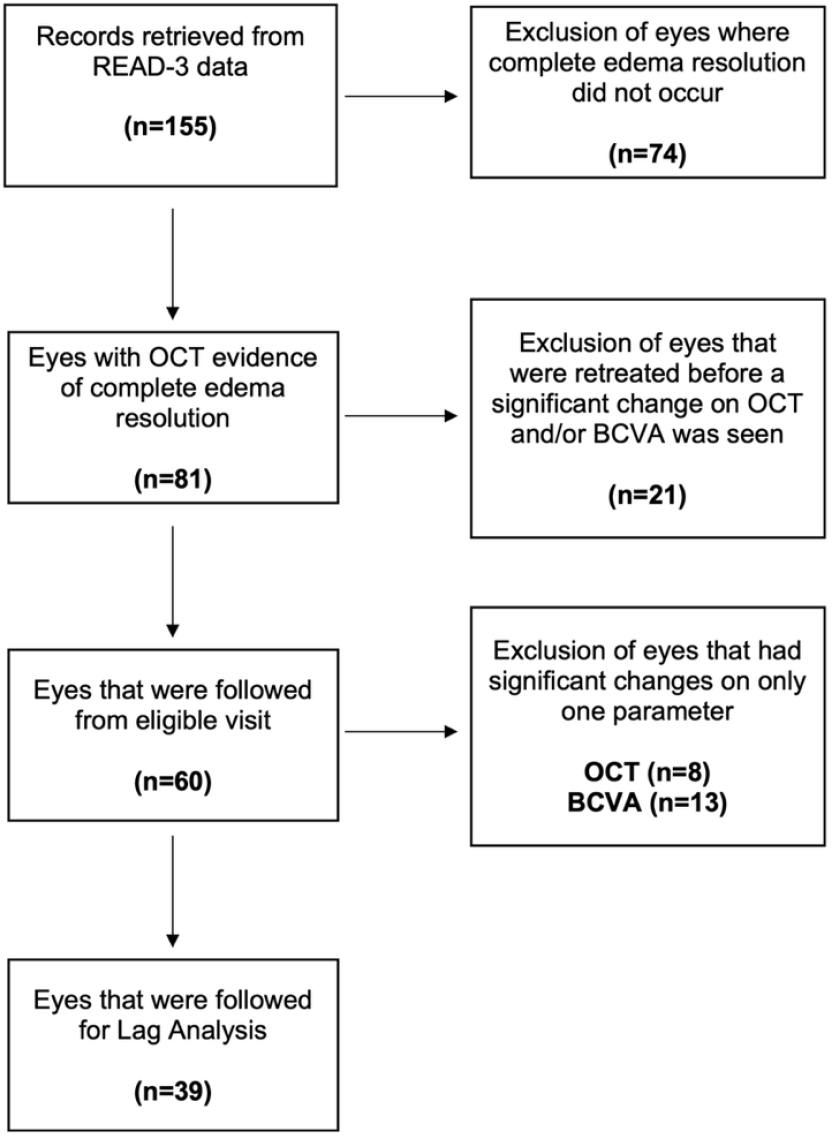
Flow diagram illustrating the identification of eligible eyes for inclusion in this analysis.

### Patient Characteristics and Disease Activity

The 60 patients had a mean age of 65.0 ± 11.0 years; 48.3% were female (29/60). All had type 1 or type 2 diabetes with DME and had received treatment as per protocol. At baseline, the mean CRT was elevated (413.9 ± 126.8 µm) and BCVA was moderately impaired (58.6 ± 10.4). At eligible visit, mean CRT improved substantially (196.9 ± 30.6 µm) and BCVA improved by ∼8 letters (66.1 ± 13.8). Mean BCVA and CRT values can be found in **Table 1**. All 60 eyes had CRT <250 µm at eligibility, confirming complete edema resolution. The median time to eligibility was 10 months (IQR 6-15.5) and the median number of injections prior to eligibility was 8.5 (IQR: 6-13.5), including the six mandatory injections as per protocol.

**Table 1.** Summary of mean BCVA and CRT at 3 key study intervals: Baseline (month 0), Eligibility visit (month 6-22), Last Study Visit (month 11-24)

|  | Baseline | Eligibility Visit | Last Study Visit |
| --- | --- | --- | --- |
| Mean BCVA, (letters) | $58.6 \pm 10.4$ | $66.1 \pm 13.8$ | $66.1 \pm 13.1$ |
| Mean CRT ($\mu\text{m}$) | $413.9 \pm 126.8$ | $196.9 \pm 30.6$ | $226.5 \pm 84.4$ |
| Anatomical Success, n (%)<br>CRT $< 250 \mu\text{m}$ | 2 (3.3%) | 60 (100%) | 41 (68.3%) |

### Recurrence, Retreatment, and Clinical Management

The 60 eyes were followed monthly as per READ-3 protocol, for edema recurrence. In this cohort, functional worsening, defined as a reduction in BCVA, was the most frequent indicator, occurring in 52 eyes (86.7%). Anatomical worsening was detected by TD-OCT in 43 eyes (71.7%). 39 eyes (65.0%) demonstrated both, a reduction in BCVA and an increase in CRT on TD-OCT.

Re-injection was performed in 55 of the 60 eyes (91.7%). In the remaining five instances (8.3%), retreatment was deferred despite objective changes in OCT or BCVA based on clinical discretion. The retreated eyes received multiple additional reinjections after initial resolution, till the conclusion of READ-3’s 24-month endpoint (median number of injections = 4, IQR: 3-9.5). For the 55 eyes that were retreated, no statistically significant difference in mean BCVA was seen between BCVA at eligibility visit of edema resolution and retreatment visit (p = 0.16), or eligibility visit and final BCVA at the end of the study period (p = 0.21).

In the group of 21 eyes where a temporal relationship between anatomy and function could not be established, 13 (61.9%) had no detectable CRT changes while 8 (38.1%) had no BCVA changes before the decision to continue treatment. The treatment, a prophylactic measure, likely helped maintain a dry macula and stable vision. In those cases, we did not observe a lag because neither parameter crossed the pre-specified threshold (these eyes can be considered “censored” in time-to-event analyses).

### Anatomical-Functional Lag

The temporal relationship between structural and functional decline, illustrated in Figure 2, was analyzed in the subset of 39 eyes that experienced worsening in both parameters. In 24 eyes (61.5%), the peak anatomical worsening and functional vision loss were detected at the same visit (Lag = 0). In 9 eyes (23.1%), functional vision loss preceded anatomical recurrence (range: 1-4 months), while OCT changes preceded BCVA change in 6 eyes (15.4%), where anatomical thickening was detected 1 to 2 months (not more) prior to observed vision loss. Cumulatively, functional changes (BCVA) either preceded or coincided with anatomical recurrence in 84.6% of cases, while anatomical changes (OCT) either preceded or coincided with functional deterioration in 76.9% of cases.

**Figure 2.**
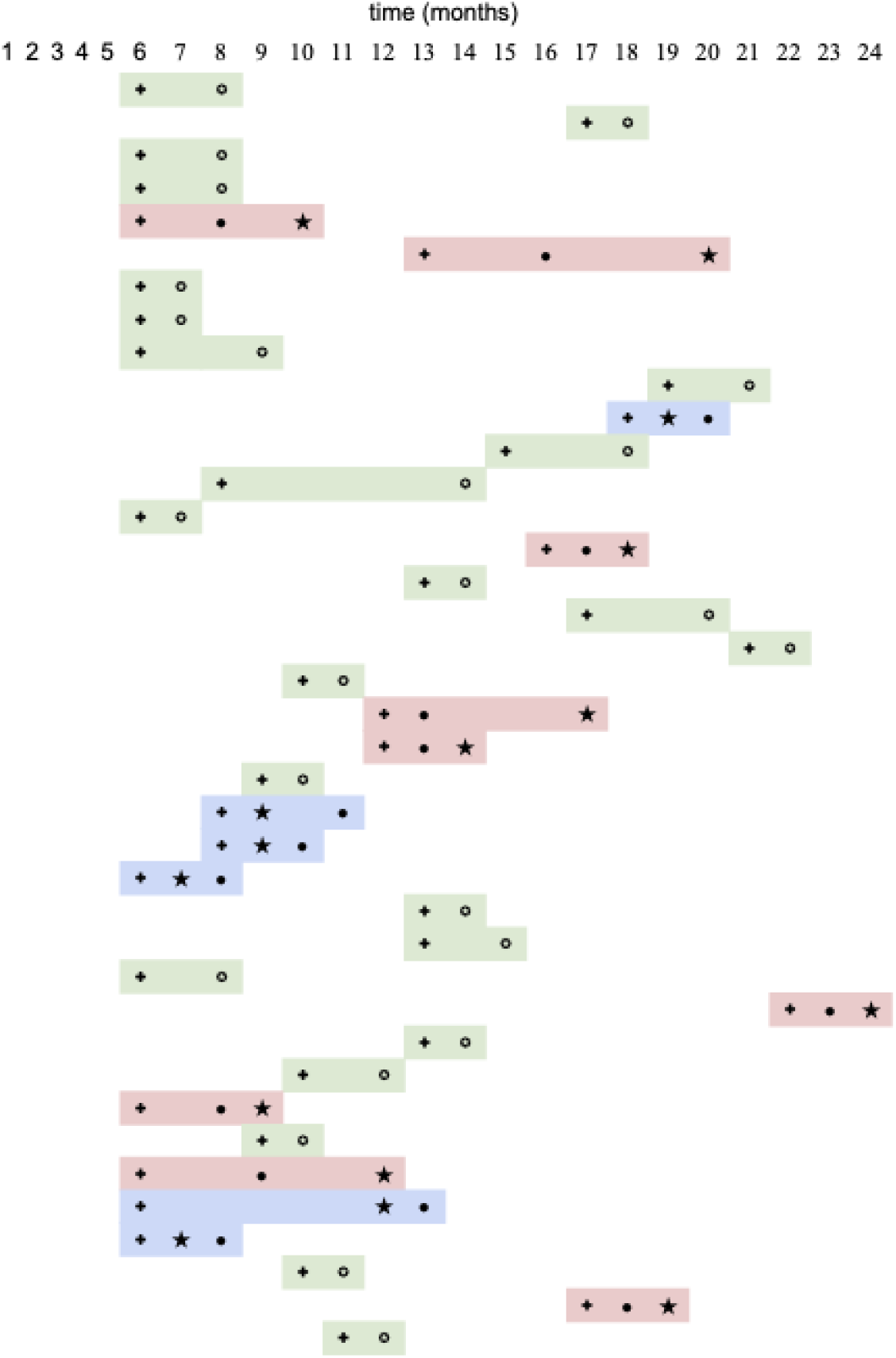
Patient-level timelines of significant BCVA and CRT changes during the analysis period. Each row represents one eye. Highlighted in green are occurrences where concurrent changes in BCVA and OCT were seen. Those highlighted in blue represent OCT-led cases, while those in red represent BCVA-led cases. Only cases with changes on both OCT and BCVA are represented. **+** eligibility visit; **★** significant change on TD-OCT (≥30 µm); ● significant BCVA change (≥4-letter); ✪ concurrent BCVA and OCT change.

## Discussion

Our post-hoc analysis of the READ-3 trial under a treat-and-extend paradigm provides intriguing evidence that BCVA monitoring can serve as an early and independent indicator of disease activity in DME. In a cohort of patients who had achieved a dry macula, we observed that BCVA changes often paralleled or even anticipated structural changes during follow-up. Notably, 23.1% (9/39) showed functional deterioration (vision loss) *preceding* any OCT-detectable fluid recurrence, in some cases by several months. This finding raises the possibility that, for select patients, declining BCVA alone could signal impending edema relapse well before OCT confirms it. To our knowledge, this is one of the first analyses to explicitly demonstrate such a lag in DME after initial successful therapy. It challenges the common assumption^10^ that “anatomical changes precede functional declines” in all cases of DME and highlights a subset where the opposite is true.

At the same time, our results reinforce that BCVA and OCT outcomes do not always move in lockstep. We saw examples of eyes maintaining good vision for a while despite OCT recurrence (CRT-led cases), as well as eyes with vision loss out of proportion to OCT findings. These scenarios underscore the multifactorial nature of DME: factors like photoreceptor damage, macular ischemia, or duration of edema can affect BCVA recovery independently of central thickness^3,11^. Prior studies have shown strong correlation between OCT and BCVA findings at baseline visits, but the correlation loses significance at subsequent study visits.^12^ Such discordances are not unique to DME; studies in retinal vein occlusion and uveitic macular edema have reported that OCT changes and BCVA gains can diverge, especially in the short term^13–16^. Our data in DME align with those reports, emphasizing the need for clinicians to monitor both aspects. In practice, improvement in CRT without commensurate BCVA gain should prompt a search for other limiting factors (e.g., retinal atrophy or diabetic cataract), while BCVA decline without OCT changes should raise suspicion for early relapse or alternative pathology (e.g., subtle vitreo-macular interface changes or capillary non-perfusion).

Crucially, because BCVA is the gold standard for visual function, these findings support a greater role for BCVA-driven monitoring in DME management. With today’s technology, if a patient-used home monitoring system detects an early BCVA drop, even a small but significant one, clinicians could intervene sooner – potentially preventing more serious edema build-up or irreversible neuroretinal damage. In our series, earlier intervention at the point of BCVA decline (rather than waiting for OCT confirmation) might have reduced the duration of edema in BCVA-leading eyes, possibly preserving a few letters of acuity that were lost. This concept dovetails with the DRCR Retina Network’s Protocol V findings^17^, where observation (no immediate treatment) in center-involved DME with good vision was found to be safe in the short term. That study suggested some eyes can tolerate edema without vision loss, but it also underscored the importance of close follow-up: patients were monitored regularly and treated if BCVA worsened. Our results extend this idea that monitoring BCVA closely is not only an option when initial vision is good, but also a potentially sensitive way to catch recurrences in a treat-and-extend scenario.

From a health systems perspective, we make the case to further study the potential of home-based BCVA monitoring, especially as we move toward longer-lasting DME treatments. While home OCT devices are being developed (e.g., home OCT for age-related macular degeneration),^18^ these are costly and require advanced technology and infrastructure. By contrast, BCVA testing needs only a reliable method to display optotypes at the correct size and distance. There are already proof-of-concept studies^19,20^ suggesting feasibility: small trials have shown that smartphone-based vision check apps can achieve near-clinic accuracy in measuring visual acuity under controlled conditions. If BCVA can indeed detect meaningful changes earlier in some patients, a reliable home test could prompt timely clinic visits or even telemedicine consultations for OCT and treatment. In effect, BCVA-based home monitoring, augmented by telemedicine, could shift care toward a more proactive and patient-centered model.

Despite proof-of-concept studies, no current app is scientifically validated or strictly adheres to ETDRS protocol. If an FDA-approved home vision test could replicate standardized 4-meter ETDRS acuity (perhaps via a smartphone app or a simple device) and securely transmit data to clinicians, it would dramatically reduce the need for patients to come in solely for monitoring. This is particularly pertinent for DME patients, who often represent a younger, working-age population struggling to balance a high individual healthcare burden. These patients frequently manage multiple comorbidities requiring visits across various clinical specialties, making the flexibility of home monitoring essential. However, rigorous validation is essential to ensure the future of home monitoring lies not in casual app use but is scientifically validated to distinguish true pathological decline from test variability.

This prioritization of functional monitoring is not only clinically prudent for treat-and-extend protocols in high-resource settings but is essential for global health equity. The simplicity and low cost of BCVA testing have particular relevance to low-and middle-income countries (LMICs), where OCT availability is limited. Prioritizing validated BCVA-based surveillance could enable earlier detection of functional decline without requiring high-cost imaging, allowing utilization of imaging resources for patients with demonstrable vision loss. We explicitly note ‘LMICs’ to highlight the global health relevance of function-first monitoring strategies. This shift promotes a more sustainable and equitable model of eye care.

The timing of monitoring is another consideration. Our data reinforce that monthly assessments are valuable – all our lag calculations were based on monthly intervals, and it was within monthly visits that these changes were captured. This aligns with the standard loading phase frequency and early extension phase in DME treatment, where monthly check-ins are common until stability is achieved. The DRCR Network has shown that after initial stabilization, follow-up intervals can be personalized (lengthened or shortened) based on disease activity^3^. Our results add that while some patients may not need monthly OCT once stable, monthly BCVA self-checks could still be prudent to detect any functional slippage between infrequent office visits. In high-resource settings, such data could refine treat-and-extend injection intervals (e.g., calling patients back sooner if home acuity drops). In lower-resource settings, monthly home BCVA monitoring could be a stopgap for those who cannot afford frequent clinic OCT – a significant public health advantage given the global diabetes epidemic.

In conclusion, while both BCVA and OCT remain invaluable for managing DME, our analysis suggests that BCVA can function as an early warning signal of disease reactivation in a significant subset of patients. Developing an accurate home-based BCVA monitoring system can transform accessible and proactive DME care. Our data support the notion that a four-week (monthly) monitoring interval is clinically meaningful for capturing the interplay between vision and retinal structure. Bridging the gap between clinic and home, BCVA-guided monitoring (supplemented by OCT as needed and expert oversight) may ultimately improve outcomes and reduce treatment burden in DME. As we move toward longer-lasting treatments and seek to globalize care, leveraging the simplicity and scalability of visual acuity measurement offers a promising avenue to ensure that no decline in vision goes unnoticed.

### Study Limitations

We acknowledge several limitations to this analysis. First, the study is a retrospective post-hoc analysis of a relatively small subgroup (n=60) from a single clinical trial. The findings, while hypothesis-generating, may not generalize to all DME patients. READ-3 had rigorous monthly follow-up and ETDRS vision testing^7^; real-world settings with less frequent visits or non-ETDRS acuity measurements might yield different results. Second, our definition of “significant” change (4 letters, 30 µm) was chosen based on practical thresholds, but various studies suggest different criteria and an interplay of multiple factors. It’s possible that micro-fluctuations in BCVA or CRT not meeting our criteria could precede larger changes. SD-OCT and Swept-Source OCT could be more sensitive in detecting anatomical changes and yield different results. Third, confounding factors could influence BCVA independent of macular edema – for example, cataract progression could cause a slow BCVA decline unrelated to retinal thickening. Additionally, factors such as the duration of disease, HbA1C levels, and age further confound.^21^ We attempted to attribute BCVA changes to DME by requiring concurrent or subsequent edema to label an event, but we cannot fully exclude other causes of vision change. Fourth, our treat-and-extend regimen was effectively monthly PRN monitoring; thus, some eyes received injections before relapse (preventing observation of a natural lag in those cases). While this reflects real clinical practice, since physicians often treat preemptively, it means we had to exclude 21 eyes as “no observable change” when in fact they might have relapsed if we had waited longer. This could bias the proportions of lag categories. A prospective study that withholds treatment until certain criteria are met might better quantify true lag frequencies but would raise ethical questions. Finally, our analysis of correlation and visual outcomes was limited by sample size, precluding definitive conclusions about long-term vision impact. The observation that vision was generally recoverable after retreatment should be interpreted cautiously; a larger dataset is needed to confirm that no harm comes from waiting for OCT changes if BCVA has already dropped.

### Implications and Future Directions

Despite these limitations, our findings support a paradigm shift wherein BCVA is not merely an outcome measure but a practical monitoring tool in DME. We envision a future DME management algorithm where patients use a certified home vision test weekly or monthly, and any confirmed BCVA decline triggers a prompt clinic evaluation (or perhaps even direct scheduling of an injection, if corroborated by other data). This could reduce unnecessary visits (for those doing well) and catch relapses sooner, optimizing treatment timing. Importantly, this approach aligns with the fact that regulatory approvals for DME therapies hinge on BCVA improvement; it makes sense to also use BCVA to guide day-to-day management, keeping the focus on what ultimately matters: the patient’s vision. With longer-acting drugs reducing visit frequency, such a scheme becomes even more attractive – with fewer clinic visits overall, and those that do occur are prompted by a real indication of change. To realize this vision, further research is needed. A prospective trial could test a BCVA-monitored strategy against an OCT-monitored strategy, measuring outcomes like number of clinic visits, injections given, vision loss prevented, and patient quality-of-life. Additionally, developing robust, user-friendly home BCVA testing technology is paramount. Integrating these tools with telemedicine platforms and electronic health records would allow seamless alerts to providers when a patient’s acuity crosses a threshold. Artificial intelligence could further assist by analyzing trends and false positives/negatives in patient-reported data.

## Conclusion

In summary, this post-hoc analysis of READ-3 highlights that clinically meaningful BCVA changes can coincide with or precede OCT changes in the management of DME. A subset of patients showed vision loss as the first sign of relapse, underscoring that BCVA is not merely a passive outcome but can be an active monitoring tool. Given practical considerations of cost, access, and patient burden, remote BCVA assessment strategy could substantially enhance DME monitoring. Our findings provide proof of concept that monthly observation of BCVA (even without OCT) can yield actionable information – potentially enabling timely retreatment and preventing avoidable vision loss. BCVA-guided decision-making, combined with clinical judgment and periodic OCT as needed, may thus represent a reliable and efficient strategy for long-term DME care. Further prospective studies are warranted to validate these findings, compare functional vs. anatomical monitoring protocols, and explore additional functional markers. By advancing vision-monitoring tools and incorporating them into treat-and-extend regimens, we can move toward a more patient-centric, scalable model of DME management that maintains vision and quality of life for millions at risk.

## Data Availability

All data produced in the present study are available upon reasonable request to the authors.

## Acknowledgments

None.

## References

1. The relationship of glycemic exposure (HbA1c) to the risk of development and progression of retinopathy in the diabetes control and complications trial. Diabetes. 1995;44(8):968–983.

2. Intravitreal Aflibercept for Diabetic Macular Edema - Ophthalmology. Accessed July 13, 2025. https://www.aaojournal.org/article/S0161-6420(14)00426-6/fulltext

3. Tang T, Tran D, Han D, Zeger SL, Crews DC, Cai CX. Place, Race, and Lapses in Diabetic Retinopathy Care. JAMA Ophthalmol. 2024;142(6):581–583. doi:10.1001/jamaophthalmol.2024.0974

4. The Relationship between OCT-measured Central Retinal Thickness and Visual Acuity in Diabetic Macular Edema. Ophthalmology. 2007;114(3):525–536. doi:10.1016/j.ophtha.2006.06.052

5. Blinder KJ, Dugel PU, Chen S, et al. Anti-VEGF treatment of diabetic macular edema in clinical practice: effectiveness and patterns of use (ECHO Study Report 1). Clin Ophthalmol Auckl NZ. 2017;11:393–401. doi:10.2147/OPTH.S128509

6. Do DV, Sepah YJ, Boyer D, et al. Month-6 primary outcomes of the READ-3 study (Ranibizumab for Edema of the mAcula in Diabetes-Protocol 3 with high dose). Eye Lond Engl. 2015;29(12):1538–1544. doi:10.1038/eye.2015.142

7. Johns Hopkins University. Ranibizumab for Edema of the Macula in Diabetes: Protocol 3 With High Dose - the READ 3 Study. clinicaltrials.gov; 2017. Accessed July 13, 2025. https://clinicaltrials.gov/study/NCT01077401

8. Early Treatment Diabetic Retinopathy Study design and baseline patient characteristics. ETDRS report number 7. Ophthalmology. 1991;98(5 Suppl):741–756. doi:10.1016/s0161-6420(13)38009-9

9. Prünte C, Fajnkuchen F, Mahmood S, et al. Ranibizumab 0.5 mg treat-and-extend regimen for diabetic macular oedema: the RETAIN study. Br J Ophthalmol. 2016;100(6):787–795. doi:10.1136/bjophthalmol-2015-307249

10. Zhang CH, Gong B, Huang C, et al. Morphological and functional changes in the macular area in diabetic macular edema after a single intravitreal injection of aflibercept. Int J Ophthalmol. 2023;16(1):88–94. doi:10.18240/ijo.2023.01.13

11. Moon BG, Um T, Lee J, Yoon YH. Correlation between Deep Capillary Plexus Perfusion and Long-Term Photoreceptor Recovery after Diabetic Macular Edema Treatment. Ophthalmol Retina. 2018;2(3):235–243. doi:10.1016/j.oret.2017.07.003

12. Diabetic Retinopathy Clinical Research Network, Browning DJ, Glassman AR, et al. Relationship between optical coherence tomography-measured central retinal thickness and visual acuity in diabetic macular edema. Ophthalmology. 2007;114(3):525–536. doi:10.1016/j.ophtha.2006.06.052

13. Yiu G, Welch RJ, Wang Y, Wang Z, Wang PW, Haskova Z. SD-OCT Predictors of Visual Outcomes after Ranibizumab Treatment for Macular Edema due to Retinal Vein Occlusion. Ophthalmol Retina. 2020;4(1):67–76. doi:10.1016/j.oret.2019.08.009

14. Sen P, Gurudas S, Ramu J, et al. Predictors of Visual Acuity Outcomes after Anti– Vascular Endothelial Growth Factor Treatment for Macular Edema Secondary to Central Retinal Vein Occlusion. Ophthalmol Retina. 2021;5(11):1115–1124. doi:10.1016/j.oret.2021.02.008

15. Iannetti L, Scarinci F, Alisi L, et al. Correlation between Morphological Characteristics of Macular Edema and Visual Acuity in Young Patients with Idiopathic Intermediate Uveitis. Medicina (Mex). 2023;59(3):529. doi:10.3390/medicina59030529

16. Matas J, Llorenç V, Fonollosa A, et al. Predictors for functional and anatomic outcomes in macular edema secondary to non-infectious uveitis. PLoS ONE. 2019;14(1):e0210799. doi:10.1371/journal.pone.0210799

17. Baker CW, Glassman AR, Beaulieu WT, et al. Effect of Initial Management With Aflibercept vs Laser Photocoagulation vs Observation on Vision Loss Among Patients With Diabetic Macular Edema Involving the Center of the Macula and Good Visual Acuity: A Randomized Clinical Trial. JAMA. 2019;321(19):1880–1894. doi:10.1001/jama.2019.5790

18. Heier JS, Liu Y, Holekamp NM, et al. Clinical Use of Home OCT Data to Manage Neovascular Age-Related Macular Degeneration. J Vitreoretin Dis. 2025;9(2):158–165. doi:10.1177/24741264241302858

19. Karampatakis V, Almaliotis D, Talimtzi P, Almpanidou S. Design and Validation of a Novel Smartphone-Based Visual Acuity Test: The K-VA Test. Ophthalmol Ther. 2023;12(3):1657–1670. doi:10.1007/s40123-023-00697-x

20. Bastawrous A, Rono HK, Livingstone IAT, et al. Development and Validation of a Smartphone-Based Visual Acuity Test (Peek Acuity) for Clinical Practice and Community-Based Fieldwork. JAMA Ophthalmol. 2015;133(8):930–937. doi:10.1001/jamaophthalmol.2015.1468

21. Bressler SB, Odia I, Maguire M, et al. Factors Associated With Visual Acuity and Central Subfield Thickness Changes When Treating Diabetic Macular Edema With Anti-Vascular Endothelial Growth Factor Therapy: An Exploratory Analysis of the Protocol T Randomized Clinical Trial. JAMA Ophthalmol. 2019;137(4):382. doi:10.1001/jamaophthalmol.2018.6786

